# Age, gender and COVID-19 infections

**DOI:** 10.1101/2020.05.24.20111765

**Authors:** Tomáš Sobotka, Zuzanna Brzozowska, Raya Muttarak, Kryštof Zeman, Vanessa di Lego

## Abstract

Data for ten European countries which provide detailed distribution of COVID-19 cases by sex and age show that among people of working age, women diagnosed with COVID-19 substantially outnumber infected men. This pattern reverses around retirement: infection rates among women fall at age 60-69, resulting in a cross-over with infection rates among men. The relative disadvantage of women peaks at ages 20-29, whereas the male disadvantage in infection rates peaks at ages 70-79. The elevated infection rates among women of working age are likely tied to their higher share in health- and care-related occupations. Our examination also suggests a link between women’s employment profiles and infection rates in prime working ages. The same factors that determine women’s higher life expectancy account for their lower fatality and higher male disadvantage at older ages.

---

Amid uncertainties around coronavirus disease 2019 (COVID-19), there is one pattern which is persistent across countries and age groups: once infected, men are more likely to die from or with COVID-19 than women^1–3^. This gender-imbalanced fatality, similar to gender disparities in mortality reported for other infectious diseases^4,5^, contrasts with a seemingly gender-neutral distribution of the confirmed COVID-19 cases: on average, men account for 49.5% cases across 40 countries with complete data on sex-specific infections and deaths^6^.

We examine whether this apparent gender equality in COVID-19 infections prevails across different age groups. COVID-19 mortality increases sharply at older ages, with very few deaths reported below age 50^7^. Therefore, age- and sex-specific pattern of infections in combination with age and sex composition of the population are key for understanding and explaining differences in COVID-19 transmission and fatality across countries^8^. We reconstruct COVID-19 infection rates by age and sex from officially reported data for ten European countries (Belgium, Czechia, Denmark, Germany, Italy, Norway, Portugal, Spain, Switzerland, and the United Kingdom (England)). This analysis reveals that the overall gender equality in total infections is achieved by a gender-unequal pattern by age that is replicated in each of the analyzed populations. Among people of working age, women diagnosed with COVID-19 substantially outnumber infected men. At older ages, this pattern reverses: there are more confirmed cases among men than among women. While this pattern has already been reported for Italy, we extend the analysis to all European countries that have published disaggregated data by age and sex^9^. The higher burden of COVID-19 infections among women of prime working ages reflects their high representation in professions that are particularly exposed to the disease.

## Women below age 60 have higher infection rates than men

We examine two sets of standardized indicators reflecting gender balance in COVID-19 infections across age groups (0-9 to 80+): reported infections rates by age and sex and the ratio of male to female infection rates (M/F ratio) by age. By using a relative measure of M/F ratio, we largely overcome the problem of cross-country differences in COVID-19 testing policies and the resulting incomparability of data on infections and infection rates.

Across all countries, cumulative infection rates are lowest (below 1 per 1000) among children and adolescents aged 0-19. On average, only 4% of all reported infections are in this age group (min. 0.6% in Spain [age 0-14], max. 9.2% in Czechia, supplement table S4). Infection rates jump to much higher level after age 20, with wide differences in age and sex profiles across countries. Whereas Portugal shows a relatively stable infection rate at ages 20-59, Czechia and Germany show an irregular profile, and Italy, Spain, and England report steeply rising infection rate with age (Figure 1). However, consistent gender differences are visible in all the analyzed countries: when compared with men, infection rates among women increase more steeply after age 20 and they remain higher throughout peak working ages, until late 50s. In Czechia, Germany, Norway and Switzerland, infection rates among women have an irregular M-shaped profile, with the peaks at ages 20-29 and 50-59 (40-49 in Czechia).

**Figure 1:**
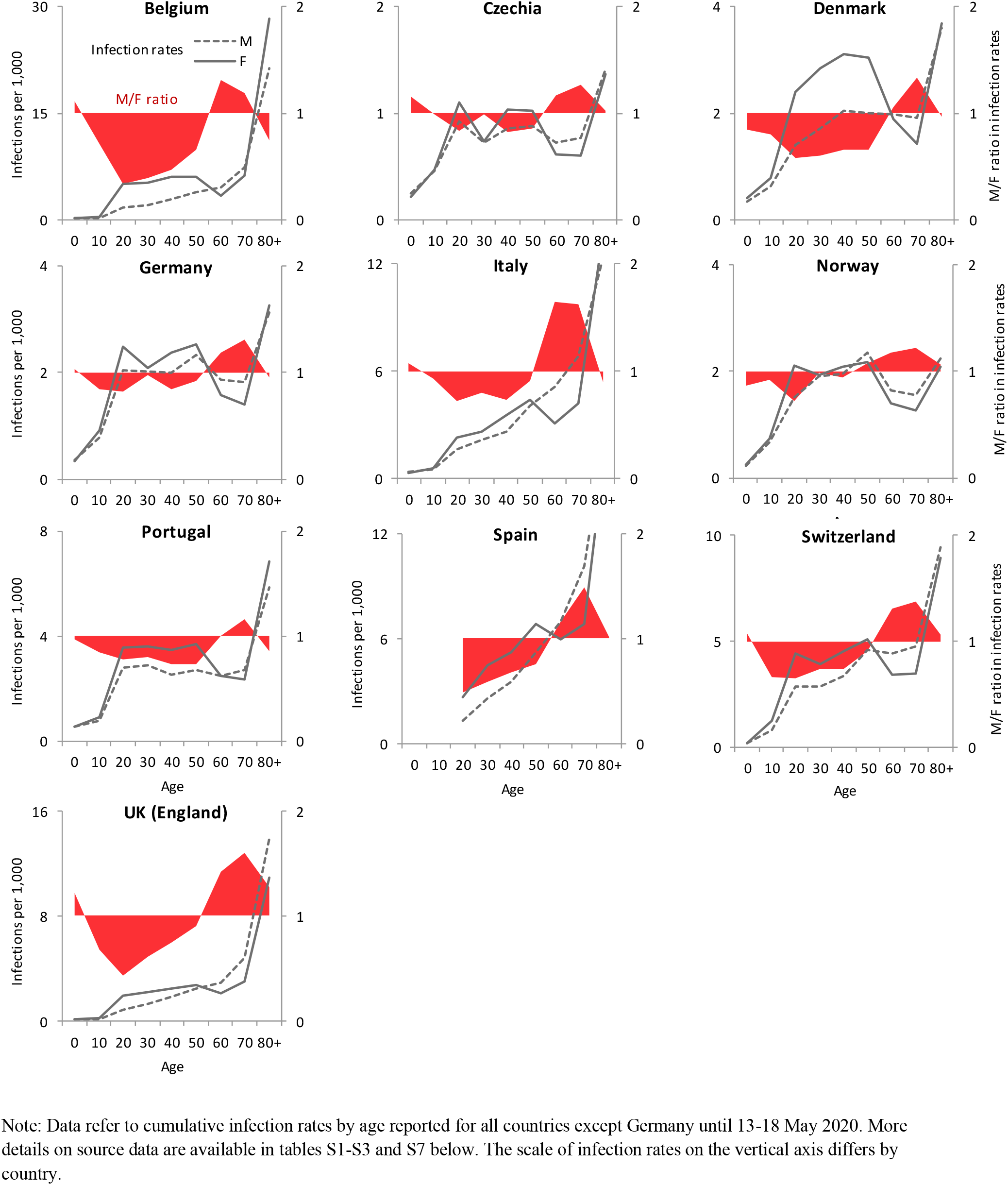
COVID-19 infection rates by age and sex per 1,000 population (solid line for females, dashed line for males, left-hand axis) and the ratio of male to female infection rates (M/F ratio) by age (red area, right-hand axis) in ten European countries

This pattern suddenly reverses after age 60, when infection rates in all countries fall steeply among women, leading to a cross-over in male and female infection rates (the cross-over in Norway occurs already after age 50). In most countries, reported infection rates for both women and men soar after age 70, most sharply in Southern Europe (Italy, Portugal and Spain), Belgium, England and Switzerland. Belgium, England, Italy and Spain also have the oldest profile of registered infections, with the mean age of infected persons surpassing 60 (60.6 in Italy to 61.6 in Italy) compared with an average of 49.5 in the other analyzed countries (table S4)..

The M/F ratios by age clearly illustrate the unequal gender profile of COVID-19 infections, already reported for Italy^9^. The female disadvantage peaks at age 20-29, when, on average, only 64 men are infected per 100 women (ranging from 34 in Belgium to 84 in Czechia) (Figure 2). In several countries, it rises again at age 40-49. Women’s dominance disappears at retirement age, between 60 and 69, when it is replaced by male overrepresentation. The male disadvantage peaks at age 70-79, when the male/female ratio implies that 136 men on average are infected per 100 women (ranging from 116 in Portugal to 162 in Italy). This pattern is not discernible when considering the absolute number of registered infections by age due to higher longevity of women and their resulting overrepresentation in the population at higher ages. Therefore, in all analyzed countries women account for most infections at ages 80+; more than one third Belgian women with reported COVID-19 infection are aged 80 or older (table S5).

**Figure 2:**
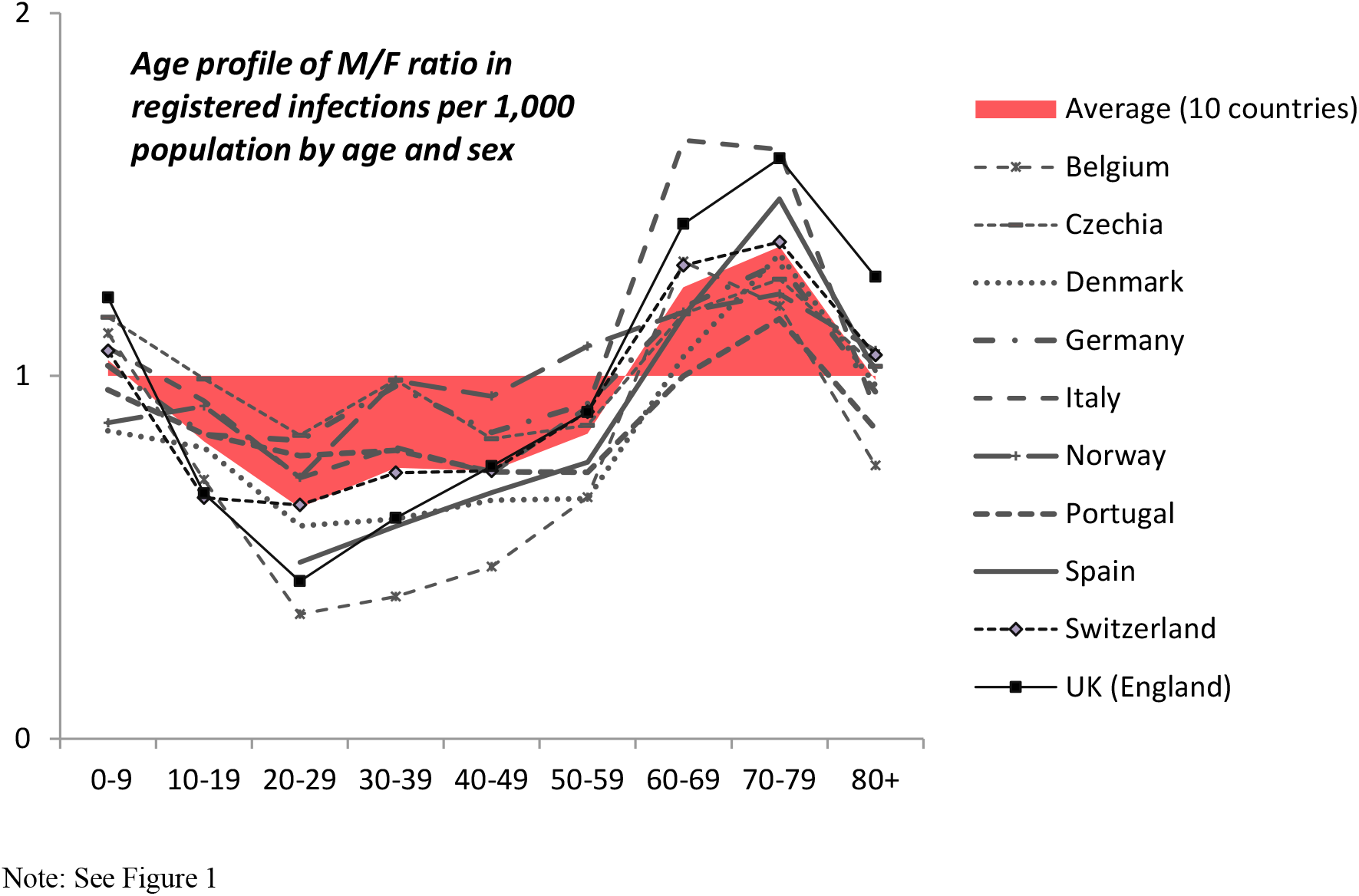
Relative ratio of male to female COVID-19 infection rates (M/F ratio) per 1,000 population by age; ten European countries

## Understanding the gender disparity in infections: the role of women’s employment and occupation status

What does this distinct age-gender pattern tell us? It clearly demonstrates the necessity of accounting for demographic factors – age and sex – in the analysis of COVID-19 pandemic^8,10^. The overall relatively balanced gender distribution in COVID-19 cases obscures starkly different age profiles of infections among men and women, which needs to be accounted for when discussing policy strategies It has been suggested that different segments of the population are better suited for returning to workplaces first given their levels of infection rates, with younger, healthier and women considered^11^. However, women of prime working age are more often diagnosed with COVID-19 than their male peers. There is no medical evidence suggesting any biological reasons for which adult women below age 60 should contract the infection more easily than men at a corresponding age. Instead, the elevated infections among women aged 20-59 are likely connected with their higher share in health- and care-related occupations: in the analyzed countries, as in other parts of Europe, between 75% and 85% of professionals in health care and social work are women^12,13^.In addition, it has been shown that health safety gear such as PPE can be inappropriate for female temple size, leaving them even more exposed to infection relative to their male counterparts. These occupations show up in the infection statistics in two ways: they are at higher risk of contracting the disease and they have a higher chance to be tested against COVID-19^14^. The drop in registered infections among women after age 60 and the overrepresentation of men among the confirmed COVID-19 cases at retirement ages give additional support to this explanation.

The “occupational disadvantage” might account for the overall higher rate of infections among women in European countries with high female labor force participation rate such as the Netherlands, Denmark and Sweden^6^. Moreover, the irregular age profile of infections among women of working age is closely correlated with their relative employment rates, which are in turn linked to their childrearing commitments as many women temporarily withdraw from employment after having a child^15^. In Czechia, Germany and Norway, where women’s employment rates—relative to male employment rates—decline in their 30s and then pick up again in their 40s, the M/F profile in infection rates follows a similar pattern (the link is weaker in Denmark) (Figure 3). In Spain and Switzerland, the relative age profile of female employment also closely matches the profile of M/F infections. The link between gender ratio of employment rates and infections is less apparent at age 60-69, when many employees have retired and employment is more selective.

**Figure 3:**
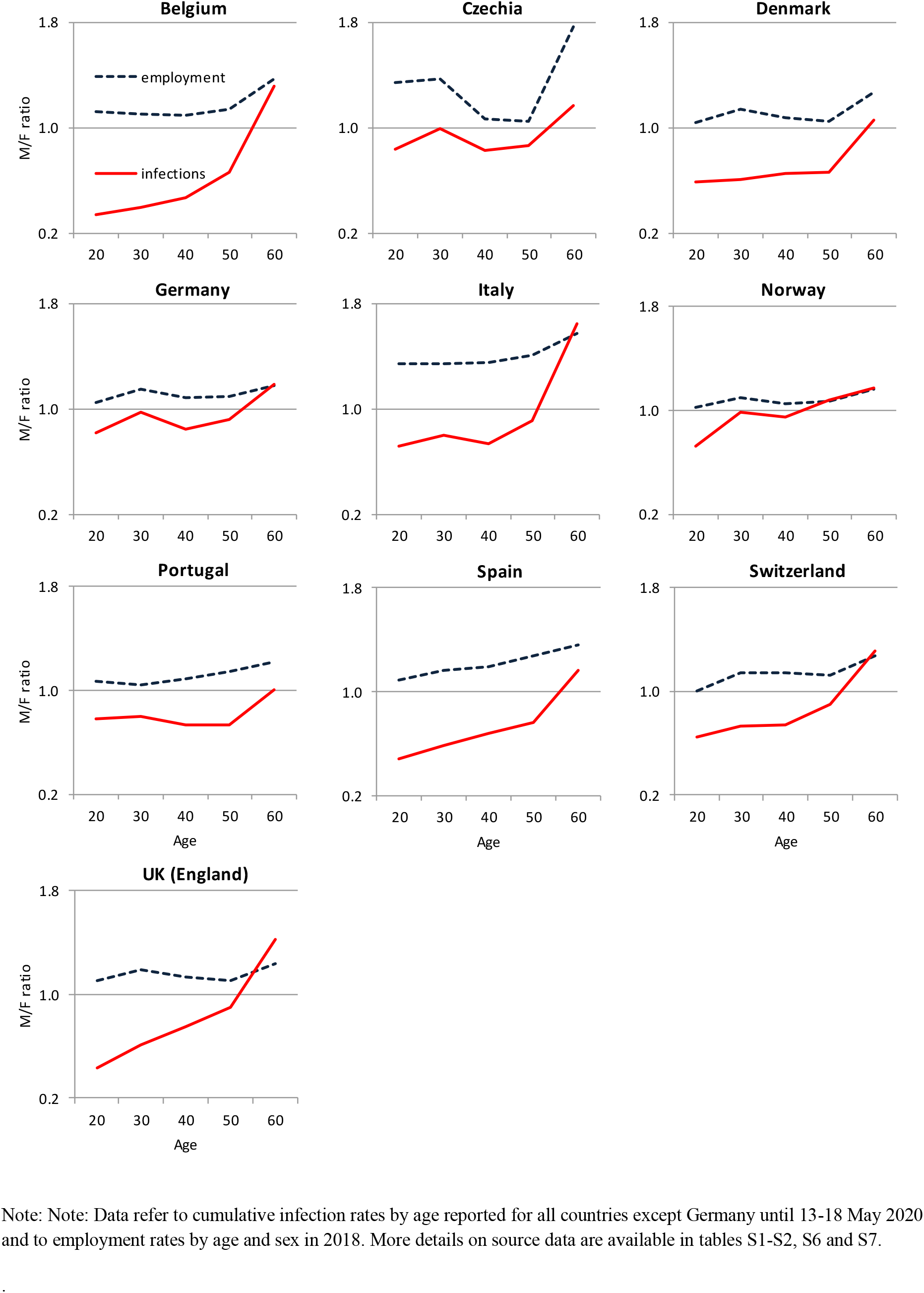
Ratio of male to female employment rates and registered COVID infections per 1,000 population, by age; ten European countries

Our analysis reveals that older men suffer from a double disadvantage: not only are they more likely to die from or with COVID-19, but the infection rate is also higher among men once they reach the most vulnerable age. More research is needed to clarify to what extent the identified age-gender disparities in the officially reported COVID-19 infections reflect the underlying distribution of infection in the population. It is possible that this disparity is affected by the selectivity of different groups to testing, with higher rates among women in health and care occupations at younger ages and among men, who are more susceptible to severe symptoms, at older ages^16,17^.

The age-specific gender differentials in COVID-19 infection reported here show that information on demographic factors need to be routinely collected to improve our understanding of vulnerability to COVID-19^8,18^. Even though the fatality rates of women below age 60 are low, engagement in care work poses higher risk to healthcare workers and care home staff. This factor should be included in the ongoing discussions on the impact of COVID-19 on women’s health and well-being^19^.

## Methods

### Data and analysis

The data on registered number of COVID-19 infections come from the official sources published by health ministries and government authorities. Mostly, these data were published as tables in regular epidemiological reports on the progression of COVID-19 or they were provided in official data repositories (Czechia, Germany). Data for Czechia were generated from a list of all individual cases. Data on employment rates among women and men in 2018 come from the OECD Family Database (see table S7 below).

Supplement tables S1-S2 provide basic characteristics of the data on reported infections. These data cover all registered cases in each country up until the reference date, which varies between 13 and 18 May for all analyzed countries except Germany (German data cover reported infections until 28 April). Most of the data were tabulated by 5- or 10-year age groups up until 80+ or 90+. Czech data specified by single years of age, whereas data for Spain showed different (and irregular) age intervals until age 29. The total registered number of infections covers over half a million of cases; in individual countries this number varies from 8.1 thousand in Czechia to 233.5 thousand in Italy. Population-wide infection rate in the analysed countries ranged from 0.76 per thousand in Czechia to 4.98 per thousand in Switzerland. Differently from a global average of the data sets reported by Global Health 5050, the data for European countries show that women dominated the statistics on COVID-19 cases, with the M/F ratio in total cases between a low of 58 men per 100 women in Belgium up to 99 men per 100 women in Norway.

Supplement table S3 summarises data on population by age and sex, which include the most recent available dataset. Except for Denmark and England, these data come from the Eurostat database and refer to 1 January 2019; data for Denmark are for 1 January 2020 and data for England refer to mid-2019. The mismatch in the reference date between population data and the data for the registered COVID-19 infections should have only a minor impact on the derived infection rates per 1,000 as the population by age and sex changes only gradually over time.

More detailed data on age and sex characteristics of persons with reported COVID-19 infections are provided in supplement tables S4-S5; source data and computed infection rates by 10-year age groups are listed in supplementary data tables in MS Excel. Supplement table S6 shows ratio of male to female employment rates by age and table S7 details all data sources.

## Data Availability

All data used in this work are official country data that are publicly available and are also explicitly detailed in the supplementary files and tables attached.

## Author’s contribution

All authors contributed to writing, data interpretation and revisions of the manuscript. T. Sobotka initiated the research, data collection, and coordinated writing and analysis. Z. Brzozowska and K. Zeman contributed to data collection, analysis and figures preparation. R. Muttarak and V. di Lego contributed to literature search.

## Conflict of interest

We declare no competing interests.

## Funding

Zuzanna Brzozowska’s work on this manuscript has been supported by Operational Programme Research, Development and Education - Project MSCAfellow2@MUNI (CZ.02.2.69/0.0/0.0/18_070/0009846).

## Acknowledgement

We acknowledge help with identifying data for Germany from Emergency Operations Centre COVID-19 of the Robert Koch Institute.

## Ethics committee approval

No approval required. Our manuscript is based on officially published data and did not involve work with individual data or patients’ records.

## SUPPLEMENT TABLES

**Tables S1 and S2:** Data on registered COVID infections, reference date, age groups format, and basic characteristics of the data sets for individual countries.

| Country | Data source | Reference date | Age groups |
| --- | --- | --- | --- |
| Belgium | 12 | 15-May-20 | 10-year, 0-9 to 90+ |
| Czechia | 1 | 14-May-20 | Single years 0 to 104 |
| Denmark | 2 | 18-May-20 | 10-year, 0-9 to 90+ |
| Germany | 10 | 28-Apr-20 | 5-year, 0-4 to 95+ |
| Italy | 3 | 14-May-20 | 10-year, 0-9 to 80+ |
| Norway | 4 | 18-May-20 | 10-year, 0-9 to 90+ |
| Portugal | 5 | 18-May-20 | 10-year, 0-9 to 80+ |
| Spain | 6 | 14-May-20 | 0-1, 2-4, 5-14, 15-29, 30-39, 10-year age groups until 80+ |
| Switzerland | 7 | 14-May-20 | 10-year, 0-9 to 80+ |
| UK (England) | 13 | 13-May-20 | 10-year, 0-9 to 80+ |

| Country | Total cases_M | Total cases_F | Total cases | Cases per 1,000 | M/F ratio (cases) |
| --- | --- | --- | --- | --- | --- |
| Belgium | 19768 | 34237 | 54005 | 4.71 | 0.58 |
| Czechia | 3858 | 4239 | 8097 | 0.76 | 0.91 |
| Denmark | 4641 | 6327 | 10968 | 1.88 | 0.73 |
| Germany | 74274 | 81570 | 155666 | 1.88 | 0.91 |
| Italy | 102628 | 119267 | 222074 | 3.68 | 0.86 |
| Norway | 4099 | 4150 | 8249 | 1.55 | 0.99 |
| Portugal | 11697 | 16622 | 28319 | 2.76 | 0.70 |
| Spain | 101373 | 133284 | 233539 | 4.98 | 0.76 |
| Switzerland | 13907 | 16528 | 30435 | 3.56 | 0.84 |
| UK (England) | 60129 | 67802 | 127931 | 2.27 | 0.89 |

Notes: Total cases may differ from the sum of male and female cases as they also include cases with unknown sex. Original data for all countries refer to absolute cases except for Switzerland, where they refer to cases per 100,000 population in a given age and gender category. Data for Switzerland also cover Liechtenstein.

**Table S3:** Data on population by age and sex.

| Country | Data source | Reference date | Age groups |
| --- | --- | --- | --- |
| Belgium | 8 | 01-Jan-19 | Single years |
| Czechia | 8 | 01-Jan-19 | Single years |
| Denmark | 9 | 01-Jan-20 | Single years |
| Germany | 8 | 01-Jan-19 | Single years |
| Italy | 8 | 01-Jan-19 | Single years |
| Norway | 8 | 01-Jan-19 | Single years |
| Portugal | 8 | 01-Jan-19 | Single years |
| Spain | 8 | 01-Jan-19 | Single years |
| Switzerland | 8 | 01-Jan-19 | Single years |
| UK (England) | 14 | mid-2019 | Single years |

**Table S4:** Mean age of women and men, and total population with reported COVID-19 infections (the last column refers to hypothetical mean age among men assuming men had the same age distribution in the population as women)

| Country | Total | Men | Women | Men (adjusted) |
| --- | --- | --- | --- | --- |
| Belgium | 60.9 | 61.9 | 60.3 | 66.4 |
| Czechia | 46.8 | 45.8 | 47.8 | 49.2 |
| Denmark | 49.1 | 50.1 | 48.3 | 52.4 |
| Germany | 50.0 | 49.3 | 50.5 | 52.3 |
| Italy | 61.6 | 60.9 | 62.1 | 64.5 |
| Norway | 46.4 | 46.8 | 46.0 | 48.5 |
| Portugal | 52.0 | 50.7 | 52.9 | 54.3 |
| Spain | 60.6 | 61.2 | 59.6 | 64.7 |
| Switzerland | 52.6 | 53.5 | 51.9 | 56.3 |
| UK (England) | 60.7 | 62.9 | 58.8 | 65.5 |
| *average* | *54.1* | *54.3* | *53.8* | *57.4* |

Note: Mean age was computed from the data grouped into 10-year age groups up to the highest category 80+ (data for Spain for ages 0-29 were grouped 0-14 and 15-29). Within each age group, we assumed that the average age corresponded to the mid-point of the age interval (e.g., 5 for age group 0-9). For the highest age group, 80+, we assumed an average of 85.

**Table S5:** Age distribution of reported COVID-19 infections (in %)

| Country | 0-19 | 20-59 | 60+ | 80+ (men) | 80+ (women) |
| --- | --- | --- | --- | --- | --- |
| Belgium | 1.6 | 46.4 | 52.0 | 25.7 | 33.7 |
| Czechia | 9.2 | 64.0 | 26.8 | 5.3 | 9.3 |
| Denmark | 6.5 | 64.0 | 29.4 | 8.4 | 9.6 |
| Germany | 5.9 | 63.9 | 30.3 | 8.5 | 13.5 |
| Italy | 2.2 | 43.9 | 53.8 | 19.4 | 31.3 |
| Norway | 7.2 | 69.1 | 23.6 | 4.8 | 7.0 |
| Portugal | 4.8 | 60.4 | 34.8 | 11.8 | 17.5 |
| Spain | 0.6 | 48.1 | 51.3 | 20.1 | 25.6 |
| Switzerland | 3.3 | 61.9 | 34.7 | 11.4 | 14.9 |
| UK (England) | 1.6 | 45.6 | 52.7 | 26.6 | 27.1 |
| *average* | *4.3* | *56.7* | *38.9* | *14.2* | *18.9* |

Note: Data for Spain are grouped into age groups 0-14 instead of 0-19 and 15-59 instead of 20-59.

**Table S6:** Ratio of male to female employment rates in 2018.

| Country | 20-29 | 30-39 | 40-49 | 50-59 | 60-69 |
| --- | --- | --- | --- | --- | --- |
| Belgium | 1.12 | 1.11 | 1.10 | 1.14 | 1.37 |
| Czechia | 1.34 | 1.37 | 1.07 | 1.05 | 1.76 |
| Denmark | 1.04 | 1.14 | 1.08 | 1.05 | 1.27 |
| Germany | 1.05 | 1.15 | 1.09 | 1.10 | 1.18 |
| Italy | 1.34 | 1.34 | 1.35 | 1.41 | 1.57 |
| Norway | 1.02 | 1.10 | 1.05 | 1.07 | 1.16 |
| Portugal | 1.07 | 1.04 | 1.09 | 1.15 | 1.22 |
| Spain | 1.09 | 1.16 | 1.19 | 1.28 | 1.36 |
| Switzerland | 1.00 | 1.14 | 1.14 | 1.12 | 1.27 |
| UK (England) | 1.10 | 1.19 | 1.13 | 1.11 | 1.24 |

Note: These data are computed from employment profiles by age and sex in OECD Family Database (reference 11 in Table S7 below)

**Table S7:** Data sources.

|  |  |
| --- | --- |
| 1 | Ministerstvo zdravotnictví ČR (Ministry of Health Czech Republic); Komenda M., Karolyi M., et al. COVID-19: Přehled aktuální situace v ČR. Onemocnění aktuálně [online]. Praha: Ministerstvo zdravotnictví ČR, 2020 [accessed 14 May 2020]. Accessed at: https://onemocneni-aktualne.mzcr.cz/covid-19 |
| 2 | Statens Serum Institut. COVID-19 i Danmark Epidemiologisk overvågningsrapport 18-05-2020. Data file uploaded 18 May 2020 at https://www.ssi.dk/aktuelt/sygdomsudbrud/coronavirus/covid-19-i-danmark-epidemiologisk-overvaagningsrapport |
| 3 | Istituto Superiore di Sanità (ISS). Epidemia COVID-19. Aggiornamento nazionale 14 maggio 2020 – ore 16:00<br>DATA PUBBLICAZIONE: 15 MAGGIO 2020. Accessed on 18 May 2020 at https://www.epicentro.iss.it/coronavirus/bollettino/Bollettino-sorveglianza-integrata-COVID-19_14-maggio-2020.pdf |
| 4 | NIPH. Norwegian Institute of Public Health. Daily reports about coronavirus disease (COVID-19). Age and gender distribution of COVID-19 cases as of 18 May 2020. Accessed 18 May 2020 at https://www.fhi.no/en/id/infectious-diseases/coronavirus/daily-reports/daily-reports-COVID19/ |
| 5 | Governo de Portugal - Ministério da Saúde. Daily report and statistics about COVID-19 ( Relatório de Situação Informação publicada diariamente às 12:00 ). Report published 15 May 2020 accessed 15 May 2020 at https://covid19.min-saude.pt/relatorio-de-situacao/ |
| 6 | RENAVE. CNE. CNM (ISCIII). Informe sobre la situación de COVID-19 en España. Informe COVID-19 nº 31. 14 de mayo de 2020. Report published 14 May 2020 accessed 18 May 2020 at https://www.isciii.es/QueHacemos/Servicios/VigilanciaSaludPublicaRENAVE/EnfermedadesTransmisibles/Documents/INFORMES/Informes%20COVID-19/Informe%20n%C2%BA%2031.%20Situaci%C3%B3n%20de%20COVID-19%20en%20Espa%C3%B1a%20a%2014%20de%20mayo%20de%202020.pdf |
| 7 | BAG (Bundesamt fuer Gesundheit): Neues Coronavirus: Situation Schweiz Schweiz und Fürstentum Liechtenstein. Data reported until 14 May 2020 (8am) accessed 15 May 2020 at https://www.bag.admin.ch/bag/fr/home/krankheiten/ausbrueche-epidemien-pandemien/aktuelle-ausbrueche-epidemien/novel-cov/situation-schweiz-und-international.html#1164290551 |
| 8 | Eurostat database. 2020. Population on 1 January by age and sex (table demo_pjan). Data accessed on 11-20 April 2020 at https://ec.europa.eu/eurostat/data/database |
| 9 | Statistics Denmark. StatBank database. Population on January 1, 2020. Accessed 11 Apr 2020 at https://www.statbank.dk/statbank5a/selectvarval/saveselections.asp |
| 10 | Bundesamt für Kartographie und Geodäsie, Robert Koch-Institut. Accessed 28 Apr 2020 at https://www.arcgis.com/home/item.html?id=f10774f1c63e40168479a1feb6c7ca74 |
| 11 | OECD Family Database, Age-employment profiles by sex, 2018. Data accessed on 7 May 2020 at http://www.oecd.org/els/family/LMF_1_4_Employment_profiles_over_life_course.xls |
| 12 | Sciensano. EPISTAT. Infectious Diseases Data Explorations & Visualizations Data accessed 15 May 2020 at https://epistat.wiv-isp.be/Covid/covid-19.html |
| 13 | Public Health England. Weekly COVID-19 surveillance report (Weekly Coronavirus Disease 2019 (COVID-19) Surveillance Report). Summary of COVID-19 surveillance. Accessed 18 May 2020 at https://www.gov.uk/government/news/weekly-covid-19-surveillance-report-published |
| 14 | Office for National Statistics (ONS): Estimates of the population for the UK, England and Wales, Scotland and Northern Ireland. Accessed 18 May 2020 at https://www.ons.gov.uk/peoplepopulationandcommunity/populationandmigration/populationestimates/datasets/populationestimatesforukenglandandwalesscotlandandnorthernireland |

## Notes

### Competing Interest Statement

The authors have declared no competing interest.

### Author Declarations

Our work deals with official and publicly available demographic data and does not require any IRB and/or ethics committee approval-.

